# A Novel NAD+ Supporting Supplement Increased NAD+ Levels, Improved Well-Being, and Alleviated Aging Symptoms: A Double-Blind, Placebo-Controlled Study

**DOI:** 10.1101/2025.05.14.25327611

**Authors:** Sarah A. Blomquist, Gregory Kelly, Sara Adães, Christopher R. D’Adamo, Abhimanyu Ardagh, Shawn Ramer, William Scuba

## Abstract

**Background:** A key factor in aging and many age-related diseases is the decline in nicotinamide adenine dinucleotide (NAD+) levels. Preclinical studies demonstrate promising results for NAD+ boosting in improving conditions of disease or aging. However, few human studies have shown meaningful improvements in physiological function or quality-of-life (QoL) with NAD+ boosting supplements.

**Objective:** This decentralized, double-blind, placebo-controlled study investigated the impact of Qualia NAD+®, a novel NAD+ supporting nutraceutical, on blood NAD+ levels and healthy aging, as assessed by QoL surveys.

**Methods:** Study participants were allocated to consume either Qualia NAD+ or placebo for 28 days. The primary outcomes were NAD+ levels that were measured at baseline and study end using a validated, self-administered, dried blood spot assay. Secondary outcomes included QoL measures that were assessed weekly. T-tests were used to compare NAD+ levels within and between study arms. Questionnaire data were compared using linear mixed-effects regression modeling.

**Results:** Sixty-three healthy adults aged 35 - 76 (n = 28 Qualia NAD+, n = 35 placebo) enrolled in the study and had their NAD+ levels assessed. Qualia NAD+ increased NAD+ levels by 67% compared to 4% in the placebo group (p < 0.001). Qualia NAD+ improved emotional well-being and vitality versus placebo at multiple timepoints (p < 0.05), while aging female symptoms improved in overall and somatic categories at day 28 versus placebo (p < 0.05). No improvements in aging symptoms were observed for males.

**Conclusion:** Qualia NAD+ increased NAD+ levels, enhanced QoL in all participants, and alleviated some aging symptoms in females compared to placebo. These findings support the need for longer-term clinical trials to clarify the sustained effects and underlying mechanisms of NAD+ supplementation on healthspan. Clinicaltrials.gov identifier: NCT06505967

## 1. Introduction

Nicotinamide adenine dinucleotide (NAD[H]) is a key coenzyme in cellular energy metabolism, driving adenosine triphosphate (ATP) production through oxidation-reduction (redox) reactions that support catabolic pathways like glycolysis, the tricarboxylic acid cycle, oxidative phosphorylation, and beta-oxidation. All paths to produce ATP require NAD+ and its reduced counterpart, NADH, and the NAD+/NADH ratio is a common point of control that links hundreds of reactions throughout a cell^1^. NAD+ is also an essential cofactor for sirtuins (SIRTs), poly-ADP-ribose polymerases (PARPs) and cyclic adenosine diphosphate (ADP)-ribose synthases (mainly ribose synthase 38 [CD38]) involved in energy regulation, inflammation and immunity, mitochondrial function, and DNA repair^2–5^. Given the critical role of NAD+ in physiology, the evidence that its levels may be reduced in humans up to 80% with increasing age and in numerous diseases has stimulated interest in the therapeutic effects of NAD+ boosting^6–13^.

In preclinical aging and disease models, encouraging improvements in physiological function and healthspan as a result of NAD+ boosting have been observed^14–22^. More than a dozen human clinical trials administering NAD+ precursors at different doses for up to 26 weeks (with one lasting two years) have consistently demonstrated their safety^23,24^. Human studies have also shown some efficacy for NAD+ boosting therapies to improve metabolic health and reduce skin cancer incidence^25–29^, however, the overall evidence for mitigating age-related diseases is limited^24,30^. Observationally, a positive correlation between cognitive capacity and plasma NAD+/NADH has been observed in centenarians^31^. Declining muscle NAD+ levels associated with aging are also linked to muscle function, while older trained athletes maintain levels comparable to those of young controls^10^. Yet, age-related variations in NAD+ levels across tissues and changes in tissue NAD+ due to boosting strategies are inconsistent, making the current functional relevance of tissue NAD+ levels unclear^24,32,33^.

Leading researchers in this field have recently acknowledged the need for improved tools, biomarkers, and clinical trial designs to better assess the potential benefits of NAD+ boosting supplements, especially in light of the poorly understood complexities of the NAD+ metabolome^24,34–36^. Furthermore, few human studies have assessed the impact of NAD+ boosting strategies for promoting healthy aging, and evidence suggesting improvements in quality of life remains sparse^24^. Despite inadequate evidence from human clinical trials on aging-related outcomes, mounting evidence suggests NAD+ status impacts multiple hallmarks of aging^37,38^, highlighting the opportunity to investigate NAD+ boosting on promoting healthspan.

Earlier clinical trials demonstrated modest increases (40 - 59%) in NAD+ levels with a daily dose of 300 mg of nicotinamide riboside (NR) or nicotinamide mononucleotide (NMN), while more significant increases typically require higher doses (1000 - 2000 mg/d)^39^. The research team hypothesized that an alternative to higher dosing of NR or NMN would be the addition of synergistic nutrients known to play a role in NAD+ production. The rationale for the development of Qualia NAD+ and testing approach was described in more detail previously^40^. In brief, emerging preclinical evidence indicates these compounds support NAD metabolism through several complementary mechanisms. Resveratrol raises cellular NAD levels through enhancement of nicotinamide phosphoribosyltransferase (NAMPT) activity, the rate-limiting step in NAD biosynthesis, as well as through adenosine triphosphate (ATP)-dependent AMP-activated protein kinase (AMPK) activation^41–43^. Magnesium is an essential cofactor in ATP-dependent steps in NAD biosynthesis and may further facilitate this process^44–46^, and is also known to influence mitochondrial redox balance by promoting NADH oxidation (regeneration of NAD+) and NADPH regeneration^47^. Caffeine enhances the activity of nicotinamide mononucleotide adenylyltransferase (NMNAT) 2 and leads to elevated NAD+ levels^48–50^, and co-administration with NR produces larger increases in NAD metabolites than either compound alone, suggesting synergistic effects^49^. Finally, B vitamins support NAD metabolism through their roles in the kynurenine pathway and energy-production processes that depend on NAD+/NADH cycling^51–53^.

In a previous clinical trial^40^, Qualia NAD+ increased whole blood NAD+ levels by an average of 74% compared to a 4% increase in the placebo group (p < 0.001) as assessed by the dried blood spot cards. Previously, we also observed significant improvements in aging symptoms for males only, possibly due to limited sample size and reduced statistical power for secondary outcomes. In this study, we adminispartered Qualia NAD+ or a placebo to healthy adults for 28 consecutive days and compared baseline measurements of whole blood NAD+ levels and quality of life survey scores to respective end values. The objective of this study was to (i) explore efficacy and reproducibility of a dietary supplement on changing NAD+ levels measured by a novel self-administered, non-invasive blood spot assay card and (ii) to assess the impact of NAD+ boosting on healthspan, evaluated by quality of life surveys.

## 2. Materials and Methods

### 2.1. Study Product

Qualia NAD+ (Qualia Life Sciences LLC, Carlsbad, CA, USA) is a dietary supplement formulated to support the maintenance and enhancement of NAD+ levels. The serving size of two dietary capsules provides the following: 0.32 mg vitamin B1, 0.4 mg vitamin B2, 254 mg of vitamin B3 (as 234 mg niacinamide [also nicotinamide] and 20 mg niacin [nicotinic acid]), 0.49 mg vitamin B6, 166 mcg folate (as dietary folate equivalents (DFE)), 1.5 mcg vitamin B12, 37.5 mcg biotin, 1.375 mg pantothenic acid, 50 mg magnesium (as Aquamin™ magnesium hydroxide from seawater [Marigot Ltd., County Cork, Ireland]), 300 mg Niagen® (nicotinamide riboside chloride from Chromadex Corp., CA, USA), 150 mg trace mineral blend (Aquamin™), 50 mg trans-resveratrol, and 28 mg caffeine from organic Coffeeberry® (Futureceuticals, USA) whole coffee fruit extract. All of these ingredients meet the criteria for dietary supplements established by the FDA, and all dosages are safe.

### 2.2. Study Design and Intervention

This study was a double-blind, placebo-controlled parallel trial. All participants were allocated by the principal investigator in a 1:1 ratio, stratified based on age and gender, to receive a 28 day supply of either Qualia NAD+ or placebo (Nu-Flow®, rice powder in a cellulose/vegetable capsule) to be taken as 2 capsules once daily in the morning, with or without food. The primary outcome was to investigate the efficacy of Qualia NAD+ in increasing NAD+ levels in blood samples at the study’s conclusion, compared to baseline and placebo. The secondary outcomes were findings from health-related quality of life questionnaires.

### 2.3. Blinding

To maintain blinding for both participants and researchers, an outside consultant, who was not involved in the research team, labeled the supplement bottles as A or B. The research team remained unaware of the bottle contents, and this information was kept confidential until the study’s completion. In case of an emergency, their treatment assignment was to be disclosed immediately.

### 2.4. Participants

Participants were recruited during June 2024 via an online recruitment platform and interested participants completed a prescreen survey to determine eligibility. Eligible participants were healthy males and females aged 40 - 65 who provided written, informed consent and a valid cell phone number for text communication. They agreed to complete all required questionnaires, records, and diaries, and to refrain from consuming supplements, energy drinks, or any other products containing any form of vitamin B3 (including niacin, niacinamide, nicotinamide riboside, or NMN) for two weeks before the baseline NAD test and throughout the study. Participants maintained their current supplement regimen, excluding those with vitamin B3. They also agreed to self-administer the NAD fingerstick test at home for both baseline and post-intervention samples and granted Qualia Life Sciences access to the results. Exclusion criteria included women who were pregnant, breastfeeding, or planning to become pregnant during the trial, individuals with known food intolerances or allergies to any ingredients in the product, and those with psychiatric conditions, neurologic disorders, endocrine disorders, cancer, or a significant cardiovascular event within the past six months. Participants taking monoimine oxidase (MAO) inhibitors, selective serotonin reuptake inhibitors (SSRIs), or other psychiatric or neurological medications, those on immunosuppressive therapy, and individuals deemed incompatible with the test protocol were also excluded. Additionally, adults lacking the capacity to provide informed consent were not eligible to participate.

### 2.5. Ethics

The study protocol was approved by the Advarra Institutional Review Board and adhered to international ethical standards for human research, including the principles outlined in the Declaration of Helsinki^54^. Informed consent was obtained from all individual participants included in the study. The Instutional Review Board (IRB)-approved study protocol (Protcol #: Pro00079070) outlined an age range of 40 - 65 years for participant inclusion. However, during the study, the age range was expanded to include individuals aged 35 - 76 to enroll a broader participant pool, which deviated from the approved protocol. This deviation was reported to the IRB in accordance with institutional guidelines. The study was registered with ClinicalTrials.gov (Identifier: NCT06505967). All authors were offered full access to the study data and reviewed and approved the final manuscript.

### 2.6. NAD+ levels testing

Whole blood NAD+ levels were assessed using a self-administered dried blood spot cards (DBS). The test was developed by Revvity (previously part of PerkinElmer). Samples were self-collected by fingerstick using a lancet on chemically coated DBS cards (Niagen+ NAD+ Test [Revvity, USA]). NAD+ assessment by DBS has been validated previously ^55,56^. Participants were offered standardized instructions and blood spot samples were collected individually by each participant, dried, and stored at room temperature (15 – 30 °C). Participants were instructed to ship their samples within 7 days of collection. Upon arrival at the laboratory, samples could receive analysis on the same day, otherwise samples were stored at ≤ − 20 °C at the laboratory until processing. The NAD+ measurements use an assay that uses an externally calibrated isotope dilution protocol, a heavy labeled internal standard, and liquid chromatography/tandem mass spectrometry (LC-MS/MS). The analyses were conducted at an independent Clinical Laboratory Improvement Amendments/College of American Pathologists (CLIA/CAP)-accredited clinical genomics and biochemical laboratory.

### 2.7. RAND-36

The 36-Item Short Form Survey (SF-36) was developed by RAND as part of the Medical Outcomes Study. Questions are grouped into eight subscales intended to assess physical functioning, bodily pain, role limitations due to physical health problems, role limitations due to personal or emotional problems, emotional well-being, social functioning, vitality, and general health perceptions. It also includes a single item that provides an indication of perceived change in health. Each item is scored on a 0 to 100 range so that the lowest and highest possible scores are 0 and 100, respectively. Scores represent the percentage of total possible scores achieved. A high score defines a more favorable health state. Participants were asked to complete this survey on a weekly basis throughout the study duration (baseline, days 7, 14, 21 and 28).

### 2.8. Aging Males Symptoms Scale

The Aging Males’ Symptoms (AMS) Scale is a standardized tool designed to assess symptoms of health-related quality of life and severity of symptoms in aging males over time^57^. The questionnaire consists of 17 items grouped into 3 categories: psychological, somatic, and sexual. An overall score is also calculated combining the 3 categories. Each item is scored on a severity scale ranging from 1 to 5, where higher scores indicate more severe symptoms. While it can be used in males of all ages, it was designed with men over 40 in mind and is used internationally for clinical research. Participants were asked to complete this survey on a weekly basis throughout the study duration (baseline, days 7, 14, 21 and 28).

### 2.9. Aging Females Symptoms Scale

The AMS scale was modified for this study to create a version that could be used with females, however most of the questions are gender-neutral and can be used for all participants. Questions 14-16 in the AMS were male-specific, therefore question 14 was modified to produce a female version and questions 15 and 16 were eliminated. The same scoring and severity scale were applied as for the AMS. Participants were asked to complete this survey on a weekly basis throughout the study duration (baseline, days 7, 14, 21 and 28).

### 2.10. Safety and Tolerability

For each day of the 28-day study, participants were asked to complete 6 questions in a self-reported diary to assess adherence and side effects. They were also asked to record how many capsules they took, the time of day they were taken, and whether the capsules were taken with food. As a baseline prior to beginning either intervention, and on a weekly basis, participants were also asked to complete a safety and tolerability survey. The areas covered were increased anxiety/worry, headache, mood changes, stomach upset, dizziness, bloating, itching, and sexual dysfunction. Possible responses for the safety and tolerability survey were: not at all, a little, a moderate amount, a lot, and a great deal, with scores corresponding from 0-4, respectively.

### 2.11. Statistical Analysis

NAD+ levels in blood and safety and tolerability data were analyzed in Python (Python Software Company, USA) using an independent t-test to compare between groups and a dependent t-test to assess changes within groups from baseline to day 28. Survey data was analyzed in R version 4.5.0 using RStudio v2026.01.0 (Posit, Boston, Massachusetts, USA) using linear regression models or non-parametric alternatives. Assumptions of linear regression were evaluated using standard diagnostic checks. A linear mixed-effects model was utilized to compare within-group and between-group differences over time compared to baseline. The model included fixed effects for group (Qualia NAD+ vs. placebo), time (baseline, days 7, 14, 21, 28), age, sex, and the interaction between group and time, as well as a random effect for participants to account for repeated measurements within individuals. The placebo group was used as the reference group. Standardized effect sizes and confidence intervals (same scale as the measured outcome) are reported for between-groups comparisons.

When violations of normality or homoscedasticity in the residuals could not be resolved through transformation, non-parametric alternatives were used. The Wilcoxon signed-rank test was applied for within-group comparisons and the Wilcoxon rank-sum test was used for between-group comparisons. To assess interactions between group, time, age (as a factor, groups were ≥50 or ≤49), and sex, while accommodating non-normal distributions, aligned rank transformation (ART) models were employed.

This study primarily utilized a modified per protocol analysis (mPP), where participant data was excluded from the analysis for the following reasons (i) the participant indicated taking <80% of the product dosage over the trial period and (ii) the data were recorded outside the designated window of time for that assessment. A per-protocol (PP) analysis was also used to examine survey results for the participants who provided baseline and end of study NAD+ levels. The sample size was estimated for the study design, with a total of 52 participants (26 per group) needed, providing 80% power to detect an effect (Cohen’s d = 0.80) with a two-tailed t-test at a 0.05 significance level. The Benjamini-Hochberg procedure was used to correct for multiple comparisons, and false discovery rate (FDR)–adjusted p-values are reported alongside standard p-values.

## 3. Results

### 3.1. Study Population

The study recruitment was conducted in June 2024 and the intervention period was conducted between June 2024 and August 2024. Out of 170 participants who volunteered for the study and were sent at-home finger stick blood testing, a group of 102 participants successfully completed the initial test to obtain baseline NAD+ measurements and were allocated to each group and instructed to take Qualia NAD+ or placebo. Sixty-three participants successfully completed both the initial test and a second NAD+ test after taking Qualia NAD+ or a placebo for 28 days (**Figure 1**). NAD+ levels data analysis is based on these 63 participants, whose ages ranged from 35 - 76 (average 54.2 years). Participants were a mix of females (n = 39) and males (n = 24). At baseline, no significant differences when comparing NAD+ levels in the Qualia NAD+ and placebo groups were observed (p = 0.49). The most common reasons for unsuccessful NAD+ measurements were: (i) 5 did not send the initial or final at-home finger stick blood kits to the lab for analysis, (ii) 15 did not provide a sufficient blood sample in the submitted test kit, and (iii) 11 returned the sample to the lab outside of the designated collection window.

**Figure 1:**
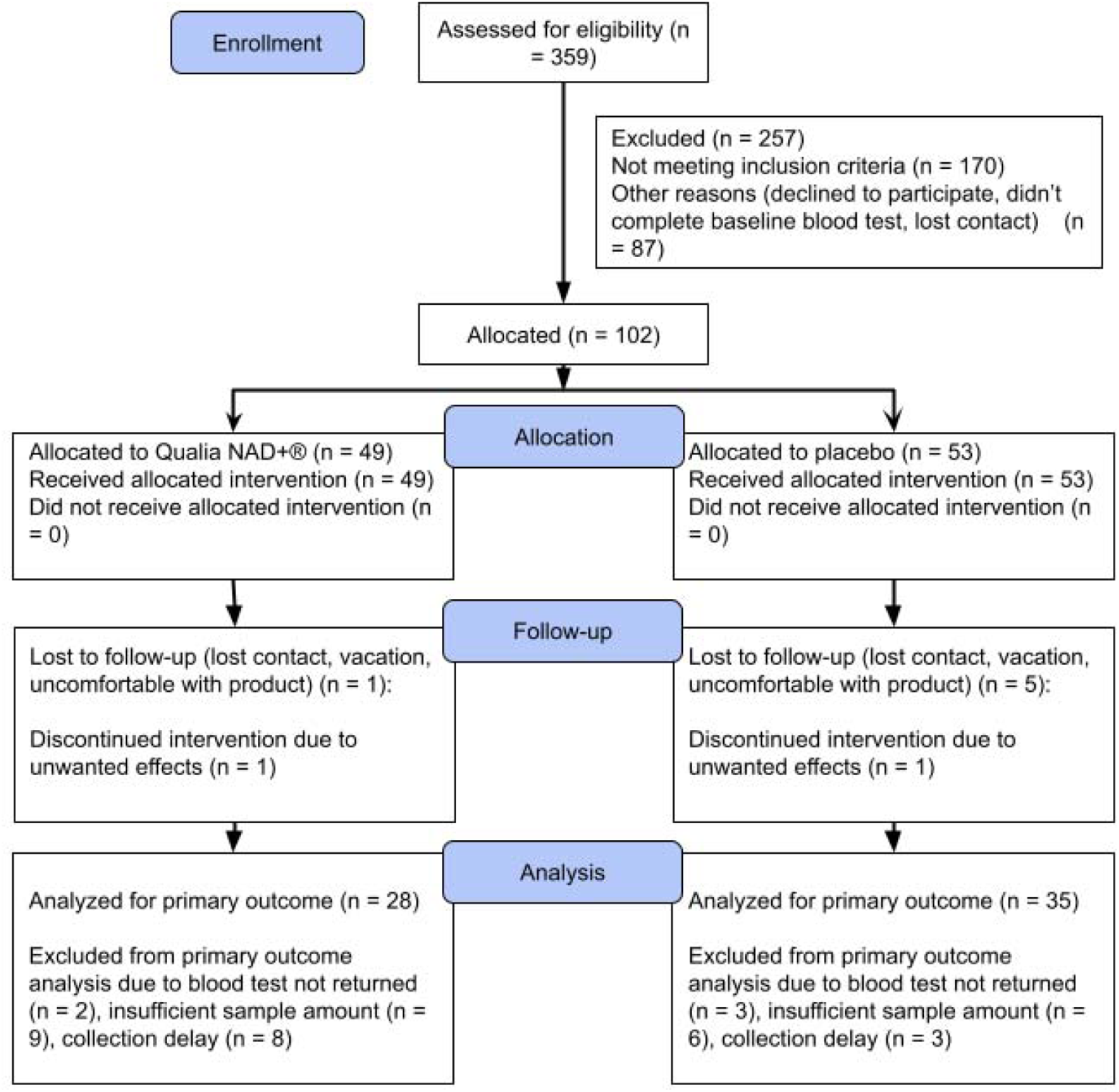
Participant Flow Diagram.

For the mPP survey analysis, 92 participants were included (n = 45 Qualia NAD+, n = 45 Placebo). Reasons for excluding participants from the mPP survey analysis are the following: (i) 20 participants did not meet the dosage threshold of 80% (described in methods), (ii) 13 participants only completed the baseline survey, and (iii) 58 participants did not complete a baseline survey. The PP survey analysis included 61 participants (n = 27 Qualia NAD+, n = 34 Placebo) who provided baseline and end of study NAD+ data and complete survey data. No significant differences in survey data at baseline were noted.

### 3.2. Primary Endpoint

Participants receiving Qualia NAD+ demonstrated a mean increase in NAD+ levels of 9.01 µM (67%) while the placebo group experienced a mean increase of 0.16 µM (4%), and these changes in scores comparing groups at the end of the study were significant (p < 0.001, Cohen’s d = 1.55, 95% CI [6.49, 12.83]) (**Table 1**). When compared to baseline values, Qualia NAD+ significantly increased NAD+ levels (p < 0.001). Significant differences in end of study NAD+ levels were also observed between-groups (Cohen’s d = 1.25, p < 0.001, 95% CI [5.28, 12.43]). Intraindividual variability was noted in response to Qualia NAD+ supplementation, with changes from baseline ranging from -62.74% to 244.41% (median 63.41%), as shown in **Figure 2**.

**Figure 2:**
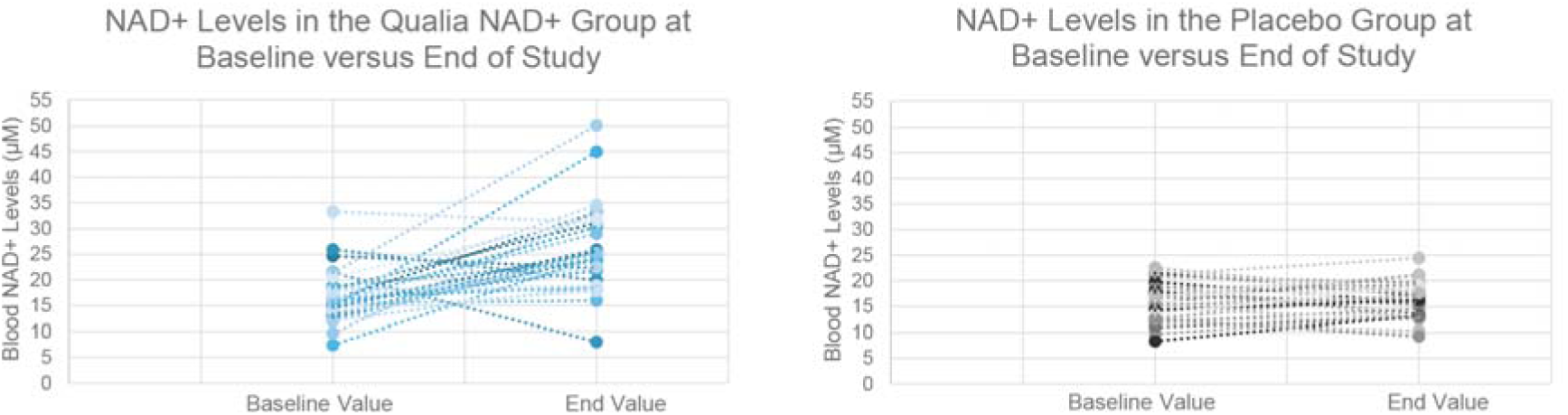
NAD+ Levels in the Qualia NAD+ and Placebo Groups. NAD values were measured using the self-administered NAD+ fingerstick at baseline (beginning of the study) and end values were measured after 28 consecutive days of supplementation in the NAD+ group (n = 28) or the placebo group (n = 35). Solid dots connected with dotted lines represent baseline and end values for an individual participant

**Table 1.** Summary Statistics for Group Comparisons. Means are shown with ± standard deviations. “Change” is the end value (day 28) minus the baseline value. * p < 0.001 when compared to the placebo group.

| Group | n | Baseline Value<br>($\mu\text{M}$) | End Value<br>($\mu\text{M}$) | Change ($\mu\text{M}$) | P value<br>(within-group) |
| --- | --- | --- | --- | --- | --- |
| NAD+ | 28 | $16.95 \pm 5.17$ | $25.95 \pm 8.70^*$ | $9.01 \pm 9.96^*$ | $< 0.001$ |
| Placebo | 35 | $16.14 \pm 3.60$ | $16.30 \pm 3.16$ | $0.16 \pm 3.24$ | 0.77 |

### 3.3. Secondary Endpoints

When comparing Qualia NAD+ to placebo using the mPP analysis, significant improvements in the “emotional well-being” category were observed for the 36-Item Short Form Survey (RAND-36) comparing baseline to days 14, 21, and 28 (**Table 2**). Improvements were also noted for the “vitality” category at day 21 and 28 compared to baseline. Using the PP analysis, significant improvements were noted in the NAD+ group versus placebo for “emotional well-being” and “vitality” comparing baseline to days 21 and 28, and in “general health” when comparing baseline to day 21 (data not shown). A trending improvement in the NAD+ group versus placebo for “general health” was also noted comparing baseline to days 14 and 28 (p = 0.08 and p = 0.06, data not shown). Nonparametric methods were used to examine the other RAND-36 categories of “physical functioning”, “role physical functioning”, “role emotional functioning”, “social functioning”, “general health”, and “pain”. No significant differences were observed between Qualia NAD+ and placebo for these categories.

**Table 2.** Probability Values for the 36-Item Short Form Survey Categories. P-values were derived from a linear mixed model with covariate adjustments for age, sex, group*time and random effects. 95% confidence intervals (CI) are reported on the same scale as the measured outcome. Effect sizes are reported as standardized beta coefficients. All p-values depicted are considered statistically significant at p < 0.05.

| Category | Day Compared to Baseline | P-Value | FDR P-Value | Effect Size | 95% CI |
| --- | --- | --- | --- | --- | --- |
| Emotional well-being | 14 | 0.005 | 0.01 | 0.28 | (1.45, 8.15) |
| Emotional well-being | 21 | 0.008 | 0.01 | 0.27 | (1.22, 7.95) |
| Emotional well-being | 28 | 0.002 | 0.004 | 0.30 | (1.93, 8.39) |
| Vitality | 21 | 0.02 | 0.04 | 0.24 | (0.94, 10.10) |
| Vitality | 28 | 0.02 | 0.04 | 0.23 | (0.86, 9.65) |

For the Aging Female Symptoms Scale (AFS) mPP analysis, significant improvements in overall and somatic scores were observed at day 28 compared to baseline for those taking Qualia NAD+ versus placebo (**Table 3**). In the PP analysis, significant improvements were also noted for those taking Qualia NAD+ versus placebo when comparing baseline to days 21 and 28 for “overall” health, and trending improvements comparing baseline to 14 for sexual health (p = 0.06) (data not shown). Psychological scores in the Qualia NAD+ group vs. placebo showed significant improvement from the linear mixed model (LMM) (p = 0.02), however, this data subset violated some LMM assumptions, and no significant differences were detected using non-parametric methods (data not shown). For the Aging Male Symptoms Scale (AMS) scores, psychological scores also showed significant improvement from the LMM at day 28 in the Qualia NAD+ group compared to placebo (p = 0.02) in both mPP and PP analyses, but similarly failed to meet assumptions for parametric analysis. Subsequent non-parametric testing showed no significant group differences (data not shown). No other significant between-groups improvements were observed for AFS or AMS scores when evaluated with Wilcoxon signed rank tests, rank sum tests, or ART methods.

**Table 3.**
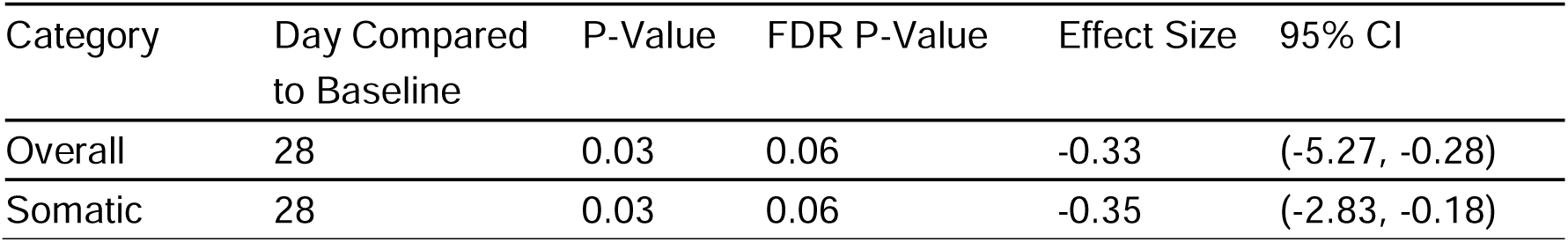
Probability Values for the Aging Female Symptom Scale. P-values were derived from a linear mixed model with covariate adjustments for group, time, group*time, age, and random effects. 95% confidence intervals (CI) are reported on the same scale as the measured outcome. Effect sizes are reported as standardized beta coefficients. All p-values depicted are considered statistically significant at p < 0.05.

### 3.4. Safety and Tolerability

There were no significant differences between NAD+ and the placebo groups in the nine safety and tolerability assessments described previously, indicating neither group experienced worsening of symptoms (p > 0.05) when comparing baseline scores to days 7, 14, 21 and 28. Overall, Qualia NAD+ was well tolerated by participants. Two participants discontinued participation due to undesired side effects. Of these, one was in the Qualia NAD+ group (reported headaches) and one was in the placebo group, revealing no significant difference in side effect related dropout between the study arms (Fisher’s exact p = 1.00).

## 4. Discussion

Since the mid-20th century, life expectancy has increased by nearly 30 years, but healthspan has not seen a comparable increase, largely due to the widespread prevalence of chronic diseases in aging populations^58^. The growing focus on healthy aging reflects the shift in interest from living longer, to living longer with improved quality of life. Current strategies for addressing the symptoms of biological aging include targeting the twelve hallmarks of aging ^59,60^. Nicotinamide adenine dinucleotide (NAD+) also plays a central role in multiple hallmarks of aging, where its reduction is a fundamental feature of the aging process, and enhancing NAD+ levels is a proposed strategy for counteracting age-related declines^37,38^.

The primary consumers of NAD+ in the body, namely sirtuins (SIRT), poly-ADP-ribose polymerases (PARPs) and cyclic ADP-ribose synthase 38 (CD38), collectively regulate nutrient-sensing, autophagy, immune function, and maintain genomic integrity and telomeres^61–69^. NAD+ boosting also improves mitochondrial function^70,71^. Maintenance of NAD+ levels are crucial for prevention of cellular senescence^72–74^, and preservation of stem cell functions^14,75,76^. In fact, nicotinamide (NAM) enhances stem cell activity through an NAD+-independent mechanism, suggesting a broader influence of B3 vitamins on the biology of aging^77^. NAD+ restoration in mice extends lifespan by modulating hallmarks of aging, raising important questions about the effects of NAD+ restoration in humans^14^. However, insufficient evidence from human clinical trials exists regarding the impact of NAD+ boosting supplements on healthspan during aging.

In this study, administering Qualia NAD+ to healthy adults aged 35 - 76 for 28 consecutive days resulted in significant increases in whole blood NAD+ levels along with improvements in quality of life parameters and symptoms of aging. The observed increases in NAD+ corroborate the data from our first placebo-controlled study, wherein NAD+ levels in the Qualia NAD+ group increased by an average of 74% compared to 4% in the placebo group (p < 0.001), also further validating the reproducibility of the self-administered blood spot assay^40^. The average NAD+ increase of 67% in this study is greater than what is reported to occur with a similar amount of nicotinamide riboside (NR) given alone^78^. This suggests a combination of NR with two additional NAD+ precursors and other ingredients designed to support the synthesis and salvaging of NAD+^40^, has additive benefits and is more effective in supporting overall NAD+ metabolism than NR given alone. One of the key factors proposed to drive NAD+ depletion with aging is decreased enzymatic activity in the salvage metabolic pathway^79^, a mechanism also targeted by the magnesium and resveratrol in Qualia NAD+^40^.

Excessive NAD+ use by major NAD-dependent enzymes is an effect that increases with age and is another key factor in NAD+ depletion^1,80–82^. The reductions in NAD+ observed (n=5) in the Qualia NAD+ treatment group may have resulted from acute injury, inflammation, or DNA damage, which activate major NAD-dependent enzymes. Since the tests were completed remotely and unsupervised by a trained individual, it’s possible human error, inconsistent timing of when samples were taken, and other sources of variance may have contributed to some of the heterogeneity observed. Heterogeneity in responses is often observed in NAD+ boosting trials^24^, while recent findings from NAD+ fingerstick assays reveal non-responders to supplementation express higher levels of primary NAD+-consuming enzymes^83^. In fact, inhibition of PARP1 and CD38 in preclinical models increased NAD+ levels and improved metabolic health^80,84,85^. Individual variations in NAD+ consuming enzyme activities may impact the effectiveness of NAD+ boosting strategies, emphasizing the importance of considering this in future clinical trials.

Beyond the variability introduced from the collection and processing of NAD+ using conventional measurement techniques^35^, differential responses to NAD+ boosting supplements have also been proposed to arise from factors such as age, sex, and the microbiome^24,35,39,83,86^. The results of this study, which revealed marked improvements in aging symptoms for females and no conclusive results for males underscores the need to also better understand sex-specific and other sources of individual differences in NAD+ metabolism and its correspondence to improvements in age-related diseases and the optimization of healthspan.

Qualia NAD+ was designed to support the broader NAD+ metabolome and multiple enzymes involved in the synthesis pathways^40^, prompting interest in the biological underpinnings of our findings. Accordingly, a primary limitation of this study is the lack of characterization of the broader NAD+ metabolome, which is increasingly recognized as crucial to understanding the relationship between NAD+ boosting supplements and health outcomes^34^. Additionally, while this study primarily utilized linear mixed modeling to analyze data, it is important to note that SF-36 scores are often heavily skewed due to ceiling or floor effects; and although non-parametric methods can better accommodate such distributions, they lack the modeling flexibility and statistical power of linear mixed models, especially for repeated measures, random effects, and covariate adjustments. Given the short-term nature of this study, it is also essential to investigate longer-term improvements in quality of life and aging-related symptoms. Overall, these limitations and the use of a specific NAD+ supplement limits the generalizability of our findings, underscoring the need for additional studies to understand the broader implications of our results.

Overall, results from this study demonstrate the efficacy of Qualia NAD+ in raising blood NAD+ levels by an average of 67%, along with corresponding improvements in emotional well-being, vitality, and symptoms of aging in female participants. Given the observed improvements in some healthspan markers, this study provides the rationale to conduct a longer-term clinical trial with an expanded biomarker panel to explore the mechanisms driving differential responses to healthspan improvement through NAD+ boosting with Qualia NAD+. A deeper understanding of NAD+ metabolism, gained through comprehensive biomarkers and clinical outcome assessments, may ultimately enhance our ability to optimize NAD+ therapies and meaningfully improve both lifespan and healthspan in aging populations.

## Data Availability

The data presented in this study are available on reasonable request from the corresponding author.

## Acknowledgements

Not applicable.

